# Using radio jingles to promote use of a family planning call center: A comparative interruptive time series analysis

**DOI:** 10.1101/2023.06.29.23292040

**Authors:** Dominique Meekers, Olaniyi Olutola, Lynn Abu Turk

**Author notes:** **Correspondence:** Dr. Dominique Meekers.

## Abstract

**Introduction:** This paper measures the effect of a radio jingle campaign promoting a toll-free family planning call center (*Honey&Banana*) in Nigeria on the number of information requests received.

**Methods:** Campaign effect is measured using a comparative interrupted time series design. The design assumes that without the radio campaign, the trend in the number of calls from the intervention group would have been parallel to that observed in the control group. The effect of the radio campaign is estimated as the difference between the actual and projected number of calls received.

**Results:** Before the radio campaign, the trend in the monthly number of requests for family planning information was nearly flat, typically averaging below 500 calls per month. After the start of the campaign, the number of calls immediately increased substantially. The analysis predicted a gain of 360 calls per month during the campaign period. However, the results show that the campaign effect was temporary, which minimal gain after the end of the campaign.

**Discussion:** Results from the comparative interrupted time series make a convincing case that the radio campaign substantially increased the demand for family planning information from the call center. However, there is no solid evidence that the radio jingle campaign had a longer-term effect on use of the call center after the radio broadcasts ended. Although short-term effects could be important if they benefited disadvantaged groups that cannot easily be reached through other means, we recommend that future campaigns be re-designed to facilitate permanent increases in call center use.

## 1 Introduction

This paper estimates the effect of a radio jingle campaign to promote a toll-free family planning call center. We use a comparative interrupted time series design that compares the number of family planning information requests that the call center received with the number that would have been expected in absence of the radio campaign. The study helps fill the gap in evidence about the effectiveness of program activities to increase use of family planning call centers.

Interrupted times series (ITS) are a valuable study design to measure the impact of interventions when a randomized controlled trial is not possible or not desirable. In public health, ITS designs have been used extensively to measure the effect of policies, programs, and health emergencies (Booth, Allen, Bray Jenkyn, Li, & Shariff, 2018; Carson et al., 2017; Cheng et al., 2016; Hategeka, Ruton, Karamouzian, Lynd, & Law, 2020; Hudson, Fielding, & Ramsay, 2019; Humphreys, Gasparrini, & Wiebe, 2017; Jandoc, Burden, Mamdani, Levesque, & Cadarette, 2015; Ramsay, Matowe, Grilli, Grimshaw, & Thomas, 2003; Siedner et al., 2020; Turner et al., 2020). However, few studies have used ITS to measure to effect of health events on family planning outcomes (Fuseini, Jarvis, Hindin, Issah, & Ankomah, 2022; Ma, Cecil, Bottle, French, & Saxena, 2020). This is surprising, considering that family planning programs often collect data that lend themselves to ITS analyses. Interrupted time series analyses work best when several pre- and post-intervention data points are available and when the intervention activity has a clearly defined starting point. Routine monitoring data from family planning programs (e.g., the monthly number of family planning clients or the number of contraceptives distributed) could be an ideal data source. Many family planning programs also implement program activities with a clear starting point (e.g., a social or mass media campaign to encourage contraceptive use). Hence, the low use of interrupted time series may be a missed opportunity to learn important lessons to inform future family planning programs.

The ITS design is a single-group study, without a comparison group. If the intervention is effective, the trend in the outcome measure is expected to exhibit an ‘interruption’ immediately or soon after the start of the intervention. This interruption may involve a sudden change in the level of the outcome measure and/or a change in the pace at which the outcome is changing (Bernal, Cummins, & Gasparrini, 2017). An important disadvantage of ITS is that the results may be confounded by other factors that also influence the outcome. The controlled interrupted time series design (CITS) extends the single-group ITS design by adding a control group that likely experienced the same confounding factors, but is not exposed to the intervention itself (Bernal, Cummins, & Gasparini, 2018; Bottomley, Scott, & Isham, 2019). CITS studies are considered strong alternatives for randomized controlled trials, and can produce comparable results (Fretheim, Soumerai, Zhang, Oxman, & Ross-Degnan, 2013; Fretheim et al., 2015; St.Clair, Cook, & Hallberg, 2014; St.Clair, Hallberg, & Cook, 2016).

One appealing feature of CITS analyses is that the graphical presentation of the results is easy to interpret (Jandoc et al., 2015; Penfold & Zhang, 2013; Wagner, Soumerai, Zhang, & Ross-Degnan, 2002). Hence, the analysis often starts with a visual inspection of the graphed results. Even without quantifying the magnitude of the intervention impact with statistical analyses, the graphed results can send a convincing message about program impact. This makes the study design particularly appealing for public health organizations that have fairly limited research capacity.

Using a CITS design, our study provides compelling evidence about the extent to which a radio campaign affected use of the *Honey&Banana* family planning call center, which will help the program make informed decisions about future mass media campaigns. To the best of our knowledge, this is the first study that assesses the potential for mass media campaigns to increase use of a family planning call center. Because of the simplicity of the study design, we recommend that other family planning organizations – particularly those with limited in-house research capacity – consider using CITS analyses to measure the effect of their social and mass media campaigns.

## 2 Background

*The Honey & Banana Connect* (H&B) program aims to provide confidential family planning information and services. H&B is accessible through a toll-free call center, social media, a website and phone apps. *H&B* promotes use of modern contraceptives, particularly long-acting reversible contraceptive methods. By dialing the toll-free number, customers can confidentially obtain family planning information from a trained agent. The call center offers family planning information in five languages (English, Hausa, Igbo, Yoruba, and Pidgin English) and callers who wish to use contraceptives can get a referral to a nearby affiliated family planning clinic.

Our study focuses on a radio jingle campaign conducted from December 2021 through March 2022. The main aim of the campaign was to increase awareness of the H&B call center and increase the number of callers requesting family planning information or a referral to a family planning provider. It was hoped that increasing awareness of the call center would permanently increase in the number of information requests. The campaign was broadcast on 28 radio stations in 12 states (Abia, Abuja FCT, Delta, Edo, Kano, Lagos, Nasarawa, Niger, Ogun, Oyo, Rivers, Sokoto). These states were selected because they had enough partner clinics to handle a potential increase in demand. The campaign used six jingles (in English, Pidgin or Hausa, depending on the location) that invited community members to contact the call center to get child spacing information or to get a referral to a family planning clinic. Radio stations broadcast the jingles approximately seven times per week.

Since campaign exposure cannot be randomized, other confounding factors (e.g., other programs) may have affected use of the call center. During the pre-intervention period H&B conducted two small campaigns that may have increased use of the call center. From April-May, H&B conducted a campaign to remind current and former users of Sayana Press injectables to renew their injection. In August, H&B celebrated the third anniversary of the call center with promotions at H&B’s network of affiliated family planning clinics, social media and website posts, and a small e-voucher campaign.

Nationwide promotion of the call center’s toll-free number takes place continuously through DKT’s contraceptive products, H&B social media and website. The website lists the toll-free number, but mostly focuses on information about contraceptive products. The toll-free number is also printed on the packaging of most DKT contraceptives. Social media postings that list the toll-free number occurred nearly daily throughout the study period. None of these channels specifically focused on the intervention period.

Other potential confounders include family planning programs implemented by the government and non-governmental organizations. The Society for Family Health and PSI Nigeria provide nationwide access to subsidized contraceptives and promote their use via mass and social media. The Nigerian Urban Reproductive Health Initiative promotes contraceptive use in Lagos, Kaduna and Oyo states (Johns Hopkins Center for Communication Programs, s.d.). These programs may have increased the demand for family planning information during our study period, but we are not aware of major program activities that coincided with the intervention period of the H&B radio campaign. Hence, it is unlikely that other family planning programs are a major confounder.

## 3 Data and methods

### 3.1.1 Study Design

We use a comparative interrupted time series designs to estimate the effect of the radio jingle campaign on the number of requests for family planning information received by the call center. We did not anticipate any lag between the start of the radio campaign and the desired increase in the number of incoming calls requesting family planning information (Bernal et al., 2017) because potential callers 1) have an existing unmet need for family planning information, and 2) are most likely to call soon after hearing the radio campaign while the call center short code is fresh in their memory. Hence, our calculations assume the intervention effect will start from December 2021 onward.

The CITS design uses an intent-to-treat approach that compares the intervention group with a control group that was not exposed to the radio campaign. If the radio campaign was effective, we would expect to see a notable increase in the demand for family planning information from callers in the broadcast region, but not elsewhere. Our assumption is that the trend in the control region is a good estimate of the secular trend in request for family planning information in absence of the radio campaign.

The accuracy of controlled interrupted time series (CITS) studies depends on the assumption that the control group is equivalent to the intervention group. If so, both groups should be exposed to the same confounding factors and therefore have parallel trends prior to the intervention (Bernal et al., 2018; Bottomley et al., 2019; Degli Esposti et al., 2021). We graph the pre-intervention trends in the intervention and control locations to verify whether the trends are parallel.

Ideally, both groups also have similar characteristics, as some subgroups may be more susceptible to the intervention or to a confounding intervention. If so, differences in the characteristics of the intervention and control group could affect how much the outcome changes (Bernal et al., 2018). Furthermore, if any characteristics that are associated with the outcomes change at a different pace in the two groups, then this would also be a confounder. Hence, checking for co-variate balance between the intervention and control group is recommended. We use recent survey data (National Population Commission (NPC)[Nigeria] & ICF, 2019) to assess whether women who live in the intervention and control areas have comparable characteristics.

After having established that intervention and control groups had similar pre-intervention characteristics and had similar pre-intervention trends, we estimated the number of calls that would have happened in absence of the radio campaign using the parallel tends assumption. Thus, we project that the intervention region would have had the exact same shape as in the control group, but with a higher starting point (i.e., we adjusted the level of the curve, but not the slope). We estimate the effect of the radio campaign by calculating the difference between the actual and projected monthly number of calls received after the intervention started.

### 3.1.2 Data source

We analyse routine data from the Customer Relations Management (CRM) software system that the call center uses to manage interactions with clients. To provide appropriate advice to callers, phone operators ask callers about the reason for the call, and whether they are already using a family planning method. If the caller consents, the agent also obtains basic socio-demographic background information. The call center agent enters the main reason for each call in the CRM, along with the type of advice or resolution offered to the caller (e.g., the type of counselling offered; whether they were referred to a clinic). This information is used to monitor the family planning needs of the call center clientele. Our analysis is based on CRM records of 12,490 calls for family planning information received from January 2021 through May 2022 (Olutola, Inyang, & Nwaogbo, 2023).

Our main outcome indicator is the monthly number of incoming calls that were related to family planning (e.g. requests for general family planning information, information about specific contraceptive methods, side-effects, etc.).

In the case of the H&B radio campaign, the intervention area consists of the broadcast area of the radio stations. Unfortunately the exact broadcast area of the different radio stations is unknown, as is the precise location of the callers. Because the radio campaign was implemented by radio stations in twelve states, we use the caller’s state of residence as a proxy. Callers who live in one of the twelve states where the radio stations are based are defined as the intervention group, and those who live elsewhere as the control group. However, it is unlikely that the participating radio stations reached the entire state. We also cannot completely rule out that the airwaves may have reached some customers in neighbouring states. Nevertheless, it seems reasonable to expect that radio campaign should predominantly affect customers who reside in one of the twelve states were the radio stations operate. We use an intent-to-treat analysis in which all callers who reside in the broadcast states are considered to be in the intervention group, irrespective of whether they were exposed to the H&B radio campaign.

### 3.1.3 Study limitations

In absence of a randomized controlled trial, it is not possible to know what would have happened without the radio campaign. However, in public health there are many instances where a randomized controlled trial is not feasible or not desirable. In such cases, use of a CITS design with a control group can provide the strongest evidence of program impact. Nevertheless, our estimates depend on the accuracy of the parallel trends assumption.

Having only two data points after the radio campaign ended limited our ability to estimate longer-term effects of the campaign. Although subsequent data points are available, they are not comparable due to launch of a large-scale e-voucher campaign.

## 4 Results

The CITS design assumes that the trend in the control group provides a good estimate of what would have happened in absence of the radio campaign. Our assumption is that the trend in the control region is a good estimate of the secular trend in request for family planning information in absence of the radio campaign. To verify the accuracy of that assumption, we assess 1) whether the two groups have similar characteristics, and 2) whether they have similar trends in the number of calls prior to the intervention.

### 4.1.1 Characteristics of the intervention and control groups

We used the 2018 Nigeria Demographic and Health Survey to compare the characteristics of women aged 15-19 in the intervention states with those in comparison states (National Population Commission (NPC)[Nigeria] & ICF, 2019). The results (see Supplementary file 1) show that women in the intervention area are more likely to live in an urban area (57% vs. 40%), to have secondary education (61% vs. 45%), be in the upper two wealth quintiles (63% vs. 34%), and to own a cell phone (65% vs. 50%). Because of these socioeconomic differences, women in the intervention area may be more likely to use the call center. Since the intervention area is based on the location of regional radio stations, the two groups also have a different ethnic composition. Nevertheless, they have a fairly similar percentage Muslims (50% vs. 56%). The two groups have a fairly similar distribution by age and marital status, but women in the intervention group are less likely to have five or more children (24% vs. 31%). There are no differences in the percentage who know at least one modern contraceptive method (92% for both), and women in the intervention group are only slightly more likely to use a modern contraceptive method (13% vs. 9%). Thus, the two groups are reasonably comparable in terms of demographics and family planning indicators. However, it is noteworthy that the intervention states have a smaller total population size (76 vs. 117 million) (National Bureau of Statistics [Nigeria], 2021).

### 4.1.2 Comparison of the pre-intervention trends

Next, we examined whether the number of family planning related calls in intervention and control group had parallel pre-intervention trends. Figure 1 shows the trends in the number of requests for family planning information from callers in the broadcast region and those elsewhere, with the intervention period shaded in green. Visual comparison of the trends *prior* to the intervention (January-November) shows that the number of calls from the broadcast region was consistently higher than the number from the other regions, despite the fact that the former has a smaller population size. This is consistent with our finding that the former has a higher socioeconomic status and better phone access. However, the trends in the two regions exhibit nearly identical patterns. During this period, the trends for the two regions followed a very similar pattern; with both showing peaks in April and August. The peak in April-May coincides with the campaign to promote the use of Sayana Press injectable contraceptives (personal communication). The peak in August coincides with the activities to celebrate the third anniversary of the call center, which included a small e-voucher campaign for free contraceptives (DKT International Nigeria, 2021). The anniversary also was also noted in the press, which may have further increased awareness of the call center (Brand Spur Media and Marketing Services, 2021; News Wings, 2021). Apart from these two temporary peaks, the monthly number of requests for family planning information showed fairly little variation during the pre-campaign period, and increased only minimally for either location. The similarity of the pre-intervention trends in the intervention and control location confirms that the study design helps control for confounding events that affect both locations, as is the case with any program activities that have a nationwide target.

**Figure 1.**
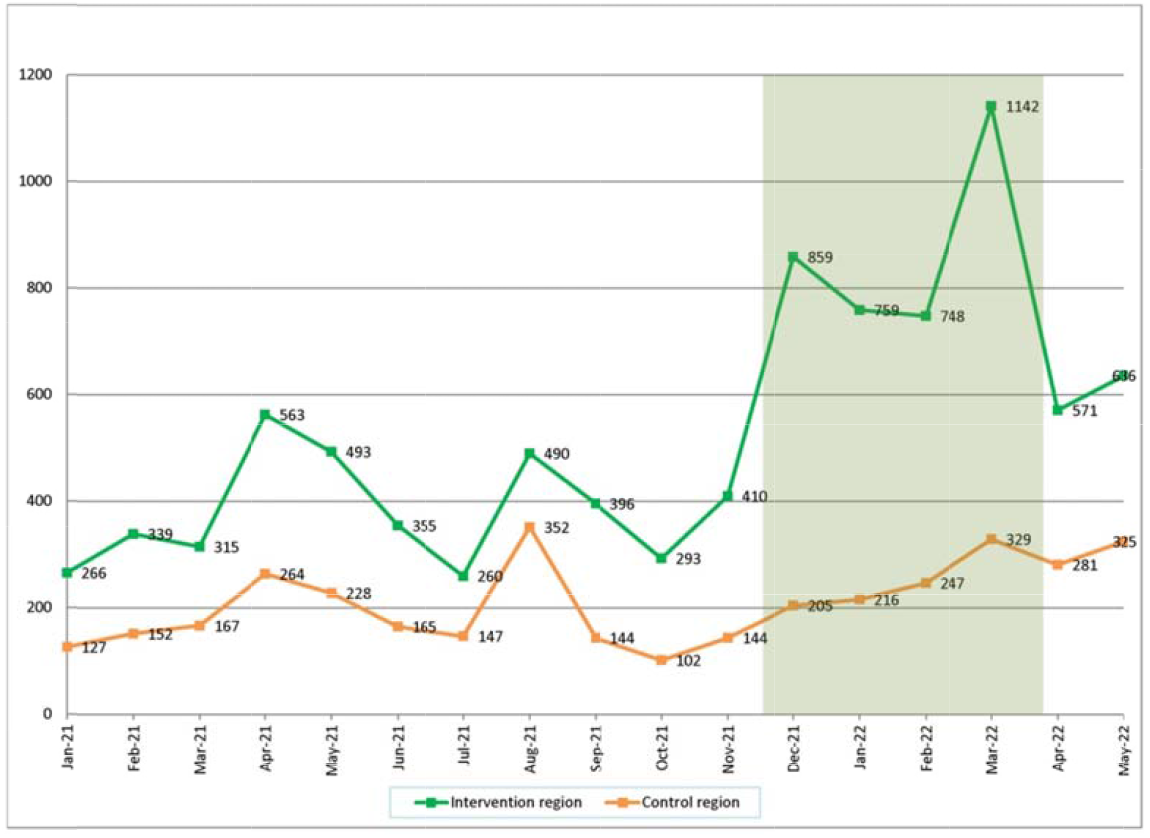
Number of requests for family planning information received, by intervention area

Calculation of the pre-intervention trend lines for the two regions confirms that the trend in the broadcast area has a much higher intercept, but only a modestly different slope. The linear trend in the broadcast region was *y* = 3.636*x* + 358.18 compared to *y* = -0.8091*x* + 185.95 for the control region. Thus, although the broadcast area started with a much higher number of calls, it increased only slightly more rapidly than in control area.

### 4.1.3 Comparison of the post-intervention trends

Upon the start of the radio campaign in December, we observe a major increase in requests for family planning information in the broadcast area, but only a steady small increase in the comparison area (Figure 1). This finding suggests that the radio campaign increased the demand for family planning information. However, it is noteworthy that the number **o**f requests for family planning information in the comparison region increased steadily between November and May, from 144 calls per month to 325. Estimation of the trend line confirms this positive slope in the number of calls (*y* =25.057*x* + 179.47).

There are two possible explanations for the observed increase in calls in the control area between November 2021 and May 2022. One possible explanation would be that the increase is caused by confounding factors, such as other program activities during this time period. Although family planning program activities by the government or other NGOs may increase the demand for family planning information, we are not aware any specific programs or activities that would have affected this specific time period. And as previously noted, neither the government nor other NGOs were directing people to the H&B call center for information. Promotion of the H&B call center is predominantly internal, by means of contraceptive product packaging, the H&B website, and H&B social media posts. However, all those H&B promotions have been going on continuously during the entire study period, without any major changes during the December-March intervention period. Hence, there are no clear known confounding factors.

A potentially more likely explanation for the increase in calls from the control area may be that there was contamination between the intervention and control area. Contamination means that some people in the control group were directly or indirectly exposed to the intervention. This may have occurred if the airwaves of the radio stations used for the campaign reached parts of a neighboring state that is located in the control area.

### 4.1.4 Estimation of the campaign effect

In Figure 2 we predict how many calls the intervention areas would have received in absence of the radio campaign. We assume that the trend in the unexposed (control) region represents the secular trend and that the two regions would have had parallel trends in absence of the radio campaign. Hence, in absence of the radio campaign the trend in the intervention region would have had the exact same shape as in the control group, but with a higher November starting point (410 calls rather than 144). The projection using the parallel trends assumption is presented by the blue line in Figure 2.

**Figure 2.**
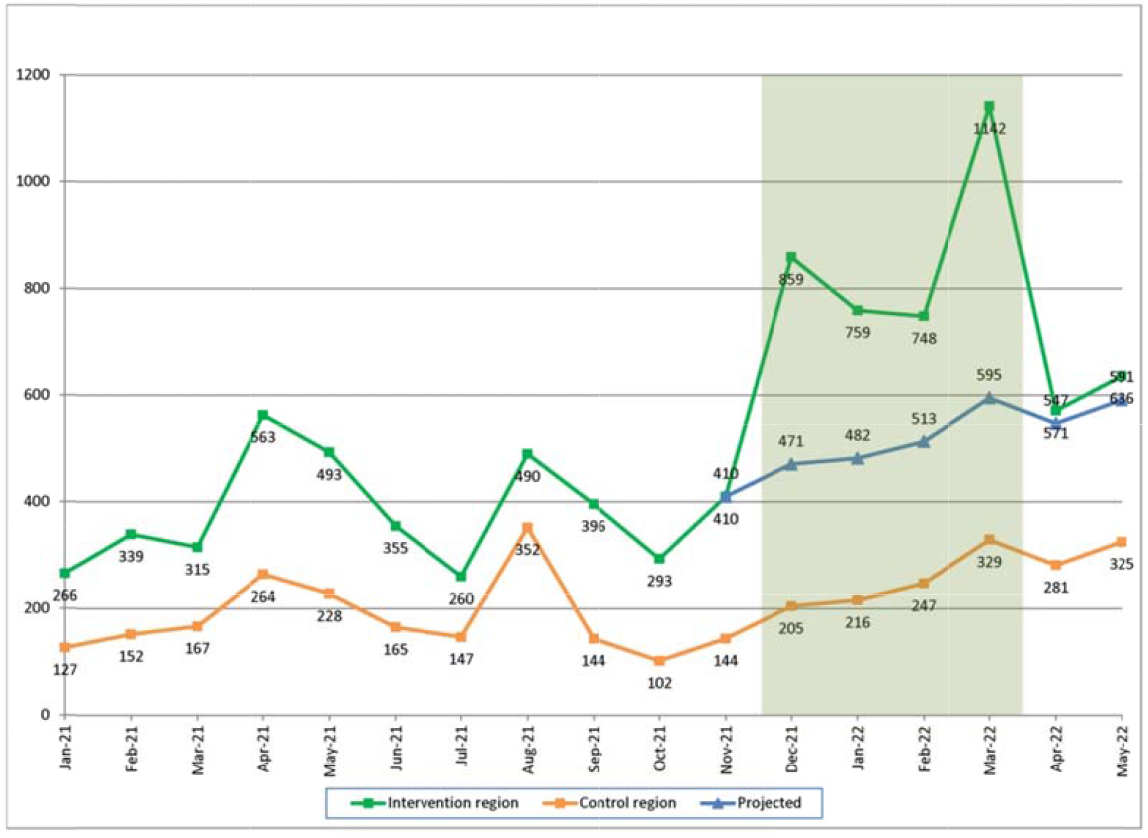
Actual and projected number of requests for family planning information, by intervention area

Comparison of the observed number of calls for the intervention region with the predicted number of calls indicates that the radio campaign resulted in a substantial increase in the number of requests for family planning information. For example, without the radio campaign the number of request for family planning information in the broadcast region is projected to have increased from 410 calls in November to only 471 in December, while the observed number increased from 410 calls to 859. This gain n the number of information requests persists for the entire duration of the radio campaign (December through March). It is noteworthy that the control group experienced a steady increase in the number of calls during the period from November 2011 through March 2022 (from 144 to 329). It is possible that the radio waves reached parts of the control area, and thereby exposed some people in the control area to the radio campaign. If so, then the parallel trends assumption will have underestimated the true effect of the intervention. However, the results for April-May 2022 show that effect of the intervention vanished completely once the campaign ended, and that the intervention and control locations once again had parallel trends. In other words, the effect of the radio campaign was only temporary.

To estimate the total number of information request gained due to the radio campaign, we calculate the difference between the actual number of calls received and the projected numbers. Table 1 shows the actual and predicted number of calls, and the estimated number of calls gained due to the campaign. For the period from December 2021 through May 2022, there was an estimated gain of only 1,515 calls. However, since some contamination of the radio campaign into the control area may have occurred, the true gain is likely to have been somewhat higher (linear projection of the pre-intervention trend in the control location would yield a gain of 1,882 calls). To put that gain in context, during the pre-intervention period the call center received roughly 500 calls per month. Hence, the added number of calls was roughly the equivalent of the volume of calls received over a 3-month period prior to the intervention. Unfortunately, the results suggest that the effect of the campaign was temporary, with negligible gain after the end of the campaign.

**Table 1:** Estimation of the number of family planning calls gained by the radio campaign (December2021 – May 2022)

|  | Control<br>group | Intervention group |  | Intervention<br>group |
| --- | --- | --- | --- | --- |
|  | Observed | Observed | Predicted* | Gained |
| 12/2 | 205 | 859 | 471 | 388 |
| 01/2 | 216 | 759 | 482 | 277 |
| 02/2 | 247 | 748 | 513 | 235 |
| 03/2 | 329 | 1,142 | 595 | 547 |
| 04/2 | 281 | 571 | 547 | 24 |
| 05/2 | 325 | 636 | 591 | 45 |
| Total | 1,603 | 4,715 | 3,119 | 1,516 |

\* CITS comparative interrupted time series results.

## 5 Discussion

This study used a controlled interrupted time series to estimate the effect of a radio jingle campaign to promote the *Honey&Banana Connect* call center on the number of requests for family planning information. The results make a convincing case that the radio campaign to increase awareness of the H&B call center generated a clear increase in the number of requests for family planning information, especially relative to the pre-campaign level of requests, but only for the duration of the campaign. In absence of strong evidence of any longer-term effects, the question becomes not whether the campaign was effective – it was – but rather whether it was worth it. The answer to that question depends largely on the original aim of the radio campaign. In the case of the H&B radio campaign, the intent was to increase awareness of the call center and its services, and to permanently increase its use. From that perspective, the campaign did not succeed. However, the aim of the H&B call center itself is to help address the need for family planning information. In terms of that larger public health objective the campaign was beneficial, as it helped address the need for family planning information for many additional customers.

Further analysis is needed to assess the extent to which the campaign benefited the most disadvantaged groups. To make informed decisions about future programs to address the need for family planning information, program managers should not only consider the cost of radio campaigns versus other potential strategies (e.g., social media, SMS messages, etc.), but also which strategies are most likely to reach disadvantaged groups with a high need for such information. In absence of evidence that the observed short-term effects benefited disadvantaged groups that cannot easily be reached through other means, we recommend that future campaigns be re-designed to facilitate permanent increases in call center use.

Our study demonstrates that simple CITS analyses can provide compelling feedback about the impact of media campaigns that can be used for programmatic decision-making. Although more advanced analytical methods exist, they require statistical expertise that is not available in many implementing public health organizations. This may discourage such organizations from using CITS analyses to inform programmatic decisions. This would be a missed opportunity, as many organizations collect routine program data that may lend themselves to CITS analyses. Our study illustrates that simple CITS analyses can provide valuable programmatic insights. We recommend that public health organizations that have limited research capacity consider conducting spreadsheet-based CITS analyses to assess the effect of their social and mass media campaigns.

## Data Availability

All data are available at Harvard Dataverse: Monthly number of phone calls received by the Honey&Banana Connect family planning call center in Nigeria (January 2021 - December 2022). https://doi.org/10.7910/DVN/OMVLLB (Olutola et al., 2023). Data are available under the terms of the Creative Commons Zero 'No rights reserved' data waiver (CC0 1.0 Public domain dedication). The Nigeria DHS data are publicly available from https://dhsprogram.com.

https://dhsprogram.com/

## Conflict of Interest

DM and OO are funded through BMGF investment INV-019286, which also funded the radio jingle campaign that is being evaluated

## Author Contributions

Meekers, D: Conceptualization, Formal Analysis, Writing – Original draft preparation; Olutola, O: Data Curation; Writing – Review & Editing; Abu Turk, L: Writing – Review & Editing; Investigation. All authors contributed to the critical revision of the manuscript and approved the final version.

## Funding

This research was conducted as part of the larger Honey and Banana Connect Project, which is funded by the Bill and Melinda Gates Foundation (investment INV-019286).

## Acknowledgments

This research was conducted as part of the Honey & Banana Connect project, which is implemented by DKT Nigeria. The authors are grateful to Mrs. Viviane Obozekhai for her feedback on an earlier draft of this paper.

## Data Availability Statement

Harvard Dataverse: Monthly number of phone calls received by the Honey&Banana Connect family planning call center in Nigeria (January 2021 – December 2022). https://doi.org/10.7910/DVN/OMVLLB (Olutola et al., 2023). Data are available under the terms of the <u>Creative Commons Zero “No rights reserved” data waiver</u> (CC0 1.0 Public domain dedication). The Nigeria DHS data are publicly available from https://dhsprogram.com.

## Consent

This study does not contain human subject data. The Tulane University Human Research Projection Office determined this study does not require IRB review and approval (Ref #2023-590)

## Notes

### Author Declarations

The Tulane University Human Research Projection Office determined this study does not require IRB review and approval (Ref #2023-590)

## References

Bernal, J. L., Cummins, S., & Gasparrini, A. (2017). Interrupted time series regression for the evaluation of public health interventions: a tutorial. Int J Epidemiol, 46(1), 348–355. doi: 10.1093/ije/dyw098

Bernal, James Lopez, Cummins, Steven, & Gasparini, Antonio. (2018). The use of controls in interrupted time series studies of public health interventions. International Journal of Epidemiology, 47(6), 2082–2093. doi: 10.1093/ije/dyy135

Booth, R. G., Allen, B. N., Bray Jenkyn, K. M., Li, L., & Shariff, S. Z. (2018). Youth Mental Health Services Utilization Rates After a Large-Scale Social Media Campaign: Population-Based Interrupted Time-Series Analysis. JMIR Ment Health, 5(2), e27. doi: 10.2196/mental.8808

Bottomley, Christian, Scott, Anthony, & Isham, Valerie. (2019). Analysing interrupted time series with a control. Epidemiologic Methods. doi: 10.1515/em-2018-0010

Brand Spur Media and Marketing Services. (2021). Celebrating 3 years of growth at Honey & Banana Connect Center. Retrieved 12/28/2022, from https://brandspurng.com/2021/09/01/celebrating-3-years-of-growth-at-honey-banana-connect-center/

Carson, K. V., Ameer, F., Sayehmiri, K., Hnin, K., van Agteren, J. E., Sayehmiri, F., … Smith, B. J. (2017). Mass media interventions for preventing smoking in young people. Cochrane Database Syst Rev, 6(6), CD001006. doi: 10.1002/14651858.CD001006.pub3

Cheng, J., Benassi, P., De Oliveira, C., Zaheer, J., Collins, M., & Kurdyak, P. (2016). Impact of a mass media mental health campaign on psychiatric emergency department visits. Can J Public Health, 107(3), e303–e311. doi: 10.17269/cjph.107.5265

Degli Esposti, M., Spreckelsen, T., Gasparrini, A., Wiebe, D. J., Bonander, C., Yakubovich, A. R., & Humphreys, D. K. (2021). Can synthetic controls improve causal inference in interrupted time series evaluations of public health interventions? Int J Epidemiol, 49(6), 2010–2020. doi: 10.1093/ije/dyaa152

DKT International Nigeria. (2021). Celebrating 3 years of growth at Honey & Banana Connect Call Center. Retrieved 12/28/2022, from https://www.youtube.com/watch?v=d5bw-LG7jWc

Fretheim, A., Soumerai, S. B., Zhang, F., Oxman, A. D., & Ross-Degnan, D. (2013). Interrupted time-series analysis yielded an effect estimate concordant with the cluster-randomized controlled trial result. J Clin Epidemiol, 66(8), 883–887. doi: 10.1016/j.jclinepi.2013.03.016

Fretheim, A., Zhang, F., Ross-Degnan, D., Oxman, A. D., Cheyne, H., Foy, R., … Soumerai, S. B. (2015). A reanalysis of cluster randomized trials showed interrupted time-series studies were valuable in health system evaluation. J Clin Epidemiol, 68(3), 324–333. doi: 10.1016/j.jclinepi.2014.10.003

Fuseini, K., Jarvis, L., Hindin, M. J., Issah, K., & Ankomah, A. (2022). Impact of COVID-19 on the Use of Emergency Contraceptives in Ghana: An Interrupted Time Series Analysis. Front Reprod Health, 4, 811429. doi: 10.3389/frph.2022.811429

Hategeka, C., Ruton, H., Karamouzian, M., Lynd, L. D., & Law, M. R. (2020). Use of interrupted time series methods in the evaluation of health system quality improvement interventions: a methodological systematic review. BMJ Glob Health, 5(10). doi: 10.1136/bmjgh-2020-003567

Hudson, J., Fielding, S., & Ramsay, C. R. (2019). Methodology and reporting characteristics of studies using interrupted time series design in healthcare. BMC Med Res Methodol, 19(1), 137. doi: 10.1186/s12874-019-0777-x

Humphreys, D. K., Gasparrini, A., & Wiebe, D. J. (2017). Evaluating the Impact of Florida’s “Stand Your Ground” Self-defense Law on Homicide and Suicide by Firearm: An Interrupted Time Series Study. JAMA Intern Med, 177(1), 44–50. doi: 10.1001/jamainternmed.2016.6811

Jandoc, R., Burden, A. M., Mamdani, M., Levesque, L. E., & Cadarette, S. M. (2015). Interrupted time series analysis in drug utilization research is increasing: systematic review and recommendations. J Clin Epidemiol, 68(8), 950–956. doi: 10.1016/j.jclinepi.2014.12.018

Johns Hopkins Center for Communication Programs. (s.d.). Nigerian Urban Reproductive Health Initiative: Using communication to increase demand for family planning. Retrieved January 1, 2023, from https://ccp.jhu.edu/projects/nigerian-urban-reproductive-health-initiative/

Ma, R., Cecil, E., Bottle, A., French, R., & Saxena, S. (2020). Impact of a pay-for-performance scheme for long-acting reversible contraceptive (LARC) advice on contraceptive uptake and abortion in British primary care: An interrupted time series study. PLoS Med, 17(9), e1003333. doi: 10.1371/journal.pmed.1003333

National Bureau of Statistics [Nigeria]. (2021). Demographic Statistics Bulletin 2021. Abuja, Nigeria: National Bureau of Statistics [Nigeria].

National Population Commission (NPC)[Nigeria], & ICF. (2019). Nigeria demographic and health survey 2018. Abuja, Nigeria and Rockville, Maryland, USA: National Population Commission and ICF.

News Wings. (2021). Honey&Banana Connect Call Center celebrates 3rd anniversary. Retrieved 12/28/2022, from https://newswings.com.ng/honeybanana-connect-call-center-celebrates-3rd-anniversary/

Olutola, Olaniyi, Inyang, Vivian, & Nwaogbo, Precious. (2023). Monthly number of phone calls received by the Honey&Banana Connect family planning call center in Nigeria (January 2021 - December 2022). Retrieved from: https://doi.org/10.7910/DVN/OMVLLB

Penfold, Robert, & Zhang, Fang. (2013). Use of interrupted time series analysis in evaluating health care quality improvements. Acad Pediatr, 13(6S), S38–S44.

Ramsay, C. R., Matowe, L., Grilli, R., Grimshaw, J. M., & Thomas, R. E. (2003). Interrupted time series designs in health technology assessment: lessons from two systematic reviews of behavior change strategies. Int J Technol Assess Health Care, 19(4), 613–623. doi: 10.1017/s0266462303000576

Siedner, M. J., Kraemer, J. D., Meyer, M. J., Harling, G., Mngomezulu, T., Gabela, P., … Herbst, K. (2020). Access to primary healthcare during lockdown measures for COVID-19 in rural South Africa: an interrupted time series analysis. BMJ Open, 10(10), e043763. doi: 10.1136/bmjopen-2020-043763

St. Clair, Travis, Cook, T, & Hallberg, Kelly. (2014). Examining the internal validiy and statistical precision of the comparative interrupted time series design by comparison with a randomized experiment. American Journal of Evaluation, 35, 311–327.

St. Clair, Travis, Hallberg, Kelly, & Cook Thomas D. (2016). The validity and precision of the comparative interrupted time-series design: Three within-study comparisons. Journal of Educational and Behavioral Studies, 41(3), 269–299.

Turner, S. L., Karahalios, A., Forbes, A. B., Taljaard, M., Grimshaw, J. M., Cheng, A. C., … McKenzie, J. E. (2020). Design characteristics and statistical methods used in interrupted time series studies evaluating public health interventions: a review. J Clin Epidemiol, 122, 1–11. doi: 10.1016/j.jclinepi.2020.02.006

Wagner, A. K., Soumerai, S. B., Zhang, F., & Ross-Degnan, D. (2002). Segmented regression analysis of interrupted time series studies in medication use research. J Clin Pharm Ther, 27(4), 299–309. doi: 10.1046/j.1365-2710.2002.00430.x

