## Supplementary Table 1 for "Using radio jingles to promote use of a family planning call center: A comparative interruptive time series analysis"

**Supplementary Table 1.** Characteristics of women aged 15-49 in the broadcast (intervention) and control states.

| | Broadcast area | | Non-broadcast area | |
|---|---|---|---|---|
| | % | No. | % | No. |
| **Type of place of residence** | | | | |
| Rural | 43.1 | 6,363 | 60.2 | 16,295 |
| Urban | 56.9 | 8,384 | 39.8 | 10,779 |
| **Level of education** | | | | |
| None | 27.3 | 4,019 | 39.1 | 10,584 |
| Primary | 12.2 | 1,803 | 15.6 | 4,235 |
| Secondary+ | 60.5 | 8,925 | 45.3 | 12,254 |
| **Wealth quintile** | | | | |
| Poorest | 10.3 | 1,524 | 21.0 | 5,698 |
| 2nd poorest | 12.4 | 1,836 | 22.9 | 6,209 |
| Middle | 14.4 | 2,126 | 22.5 | 6,081 |
| 2nd wealthiest | 24.8 | 3,651 | 19.7 | 5,340 |
| Wealthiest | 38.0 | 5,611 | 13.8 | 3,746 |
| **Owns mobile phone** | | | | |
| No | 35.5 | 5,237 | 49.7 | 1,3451 |
| Yes | 64.5 | 9,511 | 50.3 | 13,623 |
| **Ethnic group** | | | | |
| Yoruba | 27.1 | 3,992 | 9.0 | 2,426 |
| Igbo | 13.0 | 1,915 | 16.6 | 4,505 |
| Hausa | 26.7 | 3,931 | 31.4 | 8,514 |
| Other | 33.3 | 4,909 | 43.0 | 11,629 |
| **Islamic** | | | | |
| No | 50.3 | 7,422 | 44.4 | 12,027 |
| Yes | 49.7 | 7,325 | 55.6 | 15,047 |
| **Age group** | | | | |
| 15-24 | 33.9 | 5,004 | 38.0 | 10,280 |
| 35-34 | 33.5 | 4,944 | 31.4 | 8,489 |
| 35-49 | 32.5 | 4,800 | 30.7 | 8,305 |
| **Currently married or cohabiting** | | | | |
| No | 32.0 | 4,714 | 29.6 | 8,017 |
| Yes | 68.0 | 10,033 | 70.4 | 19,056 |
| **Children ever born** | | | | |
| <3 | 54.7 | 8,071 | 49.7 | 13,453 |
| 3 or 4 | 21.6 | 3,186 | 19.3 | 5,222 |
| 5+ | 23.7 | 3,490 | 31.0 | 8,399 |
| **Knows at least one modern contraceptive method** | | | | |
| No | 7.9 | 1,168 | 7.9 | 2,132 |
| Yes | 92.1 | 13,579 | 92.1 | 24,942 |
| **Current family planning use** | | | | |
| None/folkloric | 83.4 | 12,298 | 87.8 | 23,775 |
| Traditional | 4.0 | 590 | 2.8 | 762 |
| Modern | 12.6 | 1,859 | 9.4 | 2,537 |
| Total | 100.0 | 14,747 | 100.0 | 27,074 |

Source: 2018 Nigeria Demographic and Health Survey, women aged 15-49 only. Weighted data.
